# Mortality Attributed to COVID-19 in High-Altitude Populations

**DOI:** 10.1101/2020.06.10.20128025

**Authors:** Orison O. Woolcott, Richard N. Bergman

**Author notes:** Corresponding author: Orison O. Woolcott, MD, Cedars-Sinai Medical Center, 127 South San Vicente Blvd, AHSP A9220, Los Angeles, CA 90048, United States of America. **Sources of Support:** Self-funded. **AUTHORSHIP CONFIRMATION STATEMENT** OOW: study design, data collection, statistical analyses, data interpretation, final draft writing. RNB: study design, data interpretation, final draft writing. OOW and RNB have reviewed and approved the manuscript prior to submission. **AUTHOR DISCLOSURE STATEMENTS** OOW has nothing to disclose. RNB received grants from the NIH (DK7619 and DK29867).

## Abstract

**Background:** Since partial oxygen pressure decreases as altitude increases, environmental hypoxia could worsen COVID-19 patient’s hypoxemia. We compared COVID-19 mortality at different altitudes.

**Methods:** Retrospective analysis of population-level data on COVID-19 deaths in the U.S. (1,016 counties) and Mexico (567 municipalities). Mixed-model Poisson regression analysis of the association between altitude and COVID-19 mortality using individual-level data from 40,168 Mexican subjects with COVID-19, adjusting for multiple covariates.

**Results:** Between January 20 and April 13, 2020, mortality rates were higher in U.S. counties located at ≥2,000 m elevation vs. those located below 1,500 m (12.3 vs. 3.2 per 100,000; P<0.001). In Mexico, between March 13 and May 13, 2020, mortality rates were higher in municipalities located at ≥2,000 m vs. <1,500 m (5.3 vs. 3.9 per 100,000; P<0.001). Among Mexican subjects <65 years old, the risk of death was 36% higher in those living at ≥2,000 m vs. <1,500 m (adjusted incidence rate ratio: 1.36; 95% CI, 1.05-1.78; P=0.022). Among men, the risk of death was 31% higher at ≥2,000 m vs. <1,500 m (adjusted IRR: 1.31; 95% CI, 1.03-1.66; P=0.025). No association was found among women.

**Conclusion:** Altitude is associated with COVID-19 mortality in men younger than 65 years.

## INTRODUCTION

As of May 13, 2020, 4.17 million people around the world have been tested positive for SARS-CoV-2, the virus that causes Coronavirus Disease 2019 (COVID-19). Nearly 288,000 deaths have been attributed to COVID-19 (WHO). Severe hypoxemia is a common complication in critically ill patients infected with SARS-CoV-2 (Chen et al., 2020). Since partial oxygen pressure decreases as altitude increases, it is possible that environmental hypoxia could worsen COVID-19 patient’s hypoxemia.

Recently, it has been reported a lower absolute number of COVID-19 cases at higher altitudes in Bolivia and Tibet (Arias-Reyes et al., 2020). However, the interpretation of the previous findings are very difficult because data were not reported as rates (e.g. number of cases per 100,000 population) and comparison was performed in a reduced number of cities. Whether COVID-19 mortality rate is different in populations residing at low and high altitude remains unknown.

The first aim of the present study was to compare the mortality rates attributed to COVID-19 in populations residing at low and high altitudes nationwide in the United States and Mexico. The second aim was to determine the association between altitude and COVID-19 mortality adjusting for risk factors related to COVID-19 and potential confounders.

## METHODS

The present study consisted of two main analyses: 1) a retrospective analysis of data at the population level on all reported COVID-19 cases and deaths attributed to COVID-19 in counties or county equivalents of mainland United States and in municipalities of Mexico; 2) a retrospective analysis of data at the individual level on all confirmed cases of COVID-19 in Mexico. This second analysis was performed to specifically determine the association between altitude and COVID-19 outcomes (pneumonia, requirement for endotracheal intubation, and mortality) adjusting for risk factors related to COVID-19 and potential confounders.

This study did not require approval or exemption from the Cedars-Sinai Medical Center Institutional Review Board as it involved the analysis of publicly available de-identified data only.

### Data sources

#### United States

County-level data on COVID-19 cases and deaths between January 20 and April 13, 2020, were obtained from Newsbreak.com (News-Break), an online tracking source of COVID-19 outbreak in the U.S. that uses data from each state official health division. For verification, we compared the total number of cases and deaths across counties reported by Newsbreak.com and those reported by official sources from Colorado, the state with the larger number of counties located at high altitude in the U.S., and New York City, the region with the largest number of cases in the country. At the time of data collection, the number of COVID-19 cases and deaths reported by Newsbreak.com matched those that were reported by the Colorado Department of Public Health and Environment (covid19.colorado.gov) and the New York City Department of Health and Mental Hygiene (www1.nyc.gov).

#### Mexico

Individual-level data on confirmed cases of COVID-19 between January 8 and May 13, 2020 were obtained from the COVID-19 database available from the Secretary of Health of the Government of Mexico (www.gob.mx/salud). The total cumulative number of COVID-19 cases and deaths attributed to COVID-19 for each municipality was calculated from individual data.

Population estimates were obtained from the latest censuses or latest official projections. For each administrative division (county, county equivalent, and municipality), population density was calculated using the latest projected or census population.

### COVID-19 cases and deaths attributed to COVID-19

#### Population-level analysis – U.S

A total of 3,108 U.S. counties or county equivalents were initially eligible. Those with missing information on cases or deaths were excluded from analysis (n=882). Counties with zero deaths reported were also excluded (n=1,210). We used this approach to minimize possible underreport of deaths and have a fair comparison of COVID-19 mortality rates across counties. Importantly, 1,207 out of 1,210 counties with zero deaths had less than 200 COVID-19 cases. Thus, the number of fatalities in theory could be as low as 0.5 (and therefore not yet detected), given that COVID-19 fatality rate appears to range between 0.25% and 3.0% globally (Wilson et al., 2020). Final analysis included 1,016 counties.

#### Population-level analysis – Mexico

A total of 1,159 municipalities of Mexico were initially eligible. Mexican municipalities with zero deaths reported were excluded (n=592). All 592 municipalities with zero deaths had less than 48 cases of COVID-19. Final analysis included 567 municipalities.

#### Individual-level analysis – Mexico

A total of 40,186 confirmed cases of COVID-19 were initially eligible. Cases with missing information on residence location (state or municipality), pneumonia, requirement of endotracheal intubation, and intensive care unit (n=18) were excluded. Final analysis included 40,168 cases.

### Geographical elevation

For the purpose of this study, high altitude was defined as a geographical elevation ≥1,500 m (Woolcott et al., 2015). Altitude was grouped into three categories: 0-1,499 m, 1,500-1,999 m, and ≥2,000 m. Average elevation for each U.S. county and county equivalent was obtained from Zipcodes.com and validated using Google Earth (based on Geographic Coordinate System).

Average elevation for each municipality of Mexico was obtained from the Instituto Nacional para el Federalismo y el Desarrollo Municipal or using Google Earth for missing data.

### Statistical analyses

Data were presented as medians and interquartile ranges (IQR) unless otherwise indicated. Cases and deaths were presented as rates per 100,000 population. Kruskal-Wallis test (followed by post-hoc analysis with Dunn’s test with Bonferroni adjustment when appropriate) was used to compare variables across altitude categories. Wilcoxon rank-sum test was used to compare variables between survivors and non-survivors. Chi-squared test was used to compare proportions. In the Mexican population, multilevel mixed-effects Poisson regression analysis was used to estimate the relative risk of death attributed to COVID-19 (here calculated as the incidence rate ratio, IRR, with 95% confidence intervals, CIs) while accounting for nested data (states and municipalities) (Woolcott et al., 2016). The relative risk of death attributed to COVID-19 was adjusted for age, sex, medical history of diabetes, chronic obstructive pulmonary disease (COPD), cardiovascular disease, hypertension, obesity, and chronic kidney disease, and population density of residence location. We also evaluated the relative risk of pneumonia and requirement for endotracheal intubation as indicators of the severity of COVID-19. Since old age and male sex are risk factors linked to COVID-19 mortality (Li et al., 2020; Vincent and Taccone, 2020), we tested for a possible interaction between age and altitude and sex and altitude on the regression models for mortality, pneumonia, and endotracheal intubation. A P value less than 0.05 was considered statistically significant. All analyses were performed using Stata 14 (StataCorp LP, TX).

## RESULTS

Characteristics of the counties of U.S. and municipalities of Mexico are shown in Table 1 and Table 2, respectively.

**Table 1.**
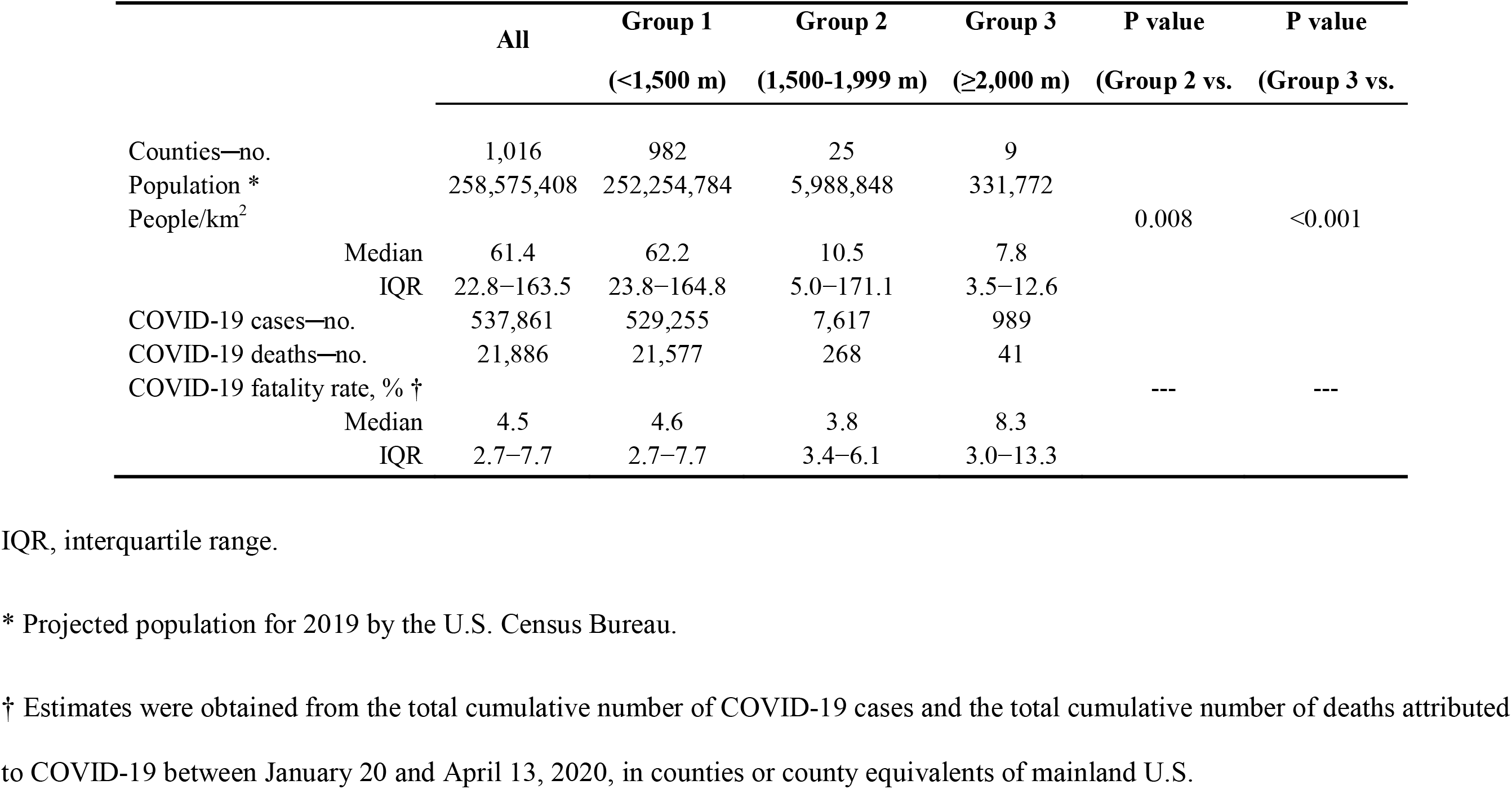
Characteristics of U.S. counties by altitude.

**Table 2.** Characteristics of municipalities of Mexico by altitude.

|  | All | Group 1<br>(<1,500 m) | Group 2<br>(1,500-1,999 m) | Group 3<br>(≥2,000 m) | P value<br>(Group 2 vs.<br>Group 1) | P value<br>(Group 3 vs.<br>Group 1) |
| --- | --- | --- | --- | --- | --- | --- |
| Municipalities—no. | 567 | 263 | 102 | 202 |  |  |
| Population * | 89,848,656 | 38,989,732 | 19,301,220 | 31,557,710 |  |  |
| People/km<sup>2</sup> |  |  |  |  | <0.001 | <0.001 |
| Median | 166.0 | 83.2 | 198.8 | 364.4 |  |  |
| IQR | 58.8–538.6 | 26.9–230.6 | 90.2–463.5 | 133.9–1,464.1 |  |  |
| COVID-19 cases—no. | 38,265 | 15,893 | 2,763 | 19,609 |  |  |
| COVID-19 deaths—no. | 4,220 | 2,099 | 302 | 1,819 |  |  |
| COVID-19 fatality rate, % † |  |  |  |  | -- | -- |
| Median | 20.0 | 20.0 | 20.0 | 20.0 |  |  |
| IQR | 10.0–40.0 | 10.0–40.4 | 12.0–50.0 | 9.5–33.3 |  |  |

IQR, interquartile range.
\* Data from latest projected population or latest censuses.
† Estimates were obtained from the total cumulative number of COVID-19 cases and the total cumulative number of deaths attributed to COVID-19 between March 13 and May 13, 2020.

### Population-level analysis – U.S

The total cumulative number of COVID-19 cases was significantly higher in U.S. counties with a mean elevation ≥2,000-m than in those below 1,500 m (176.3 vs. 67.2 per 100,000; P=0.023) (Figure 1A). COVID-19 mortality rates were also higher in counties at ≥2,000-m than in those below 1,500 m (12.3 vs. 3.2 per 100,000; P<0.001) (Figure 1B).

**Figure 1.**
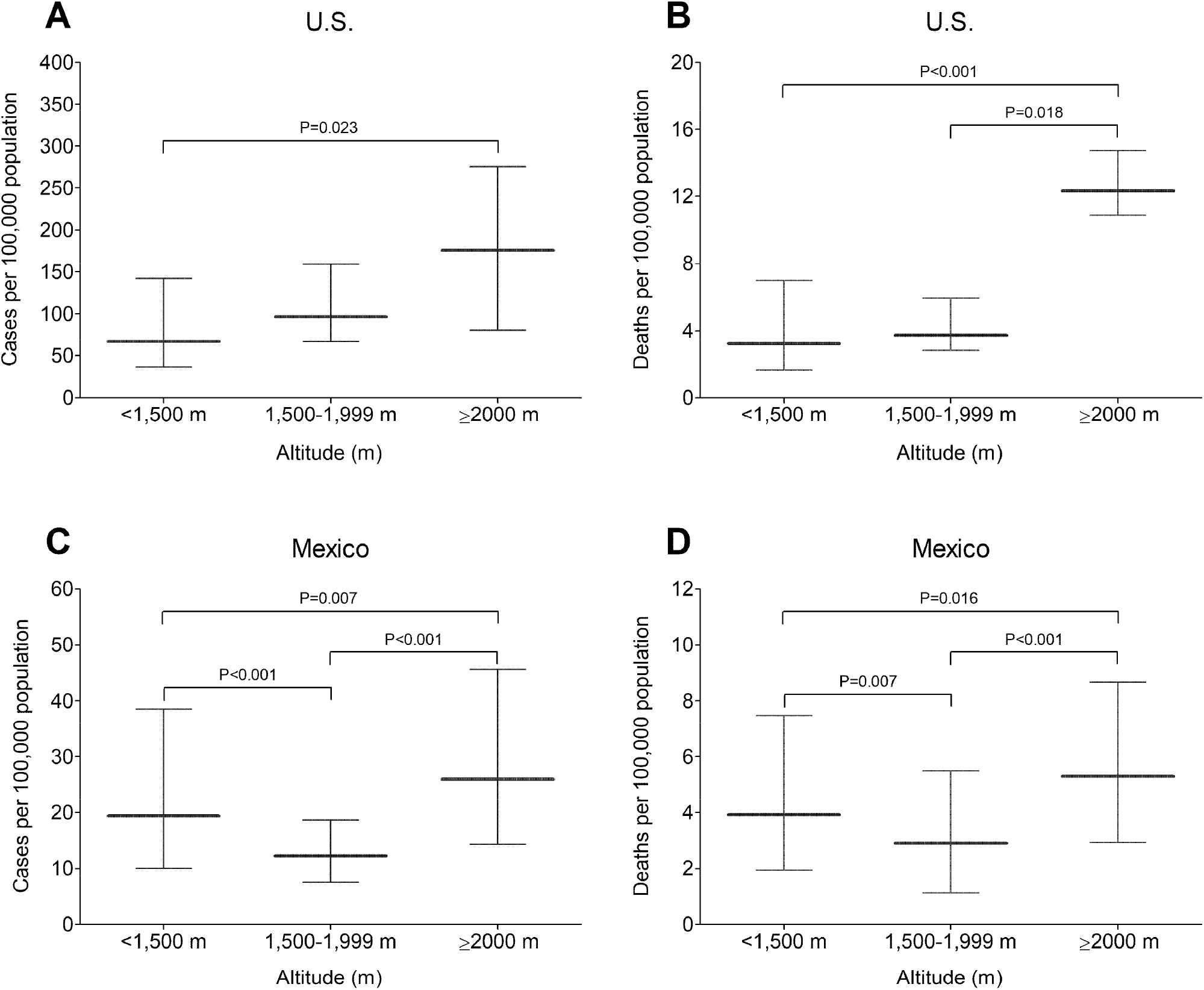
Total cumulative number of cases of COVID-19 and mortality rate attributed to COVID-19 in populations residing at high altitude. (A, B) Data comprised reported cases of COVID-19 and deaths attributed to COVID-19 between January 20 and April 13, 2020, in 1,016 counties or county equivalents of mainland U.S. grouped into three altitude categories: <1,500 m (n = 982); 1,500-1,999 m (n = 25); ≥2,000 m (n = 9). Span average altitude among U.S. counties ranged from sea level to 2,927 m. (C, D) Data comprised reported cases of COVID-19 and deaths attributed to COVID-19 up to May 13, 2020, in 567 municipalities of Mexico: <1,500 m (n = 263); 1,500-1,999 m (n = 102); ≥2,000 m (n = 202). Span average altitude among Mexican municipalities ranged from sea level to 2,905 m. Horizontal lines represent medians; vertical bars represent interquartile ranges. Kruskal-Wallis test (followed by post-hoc analysis with Dunn’s test with Bonferroni adjustment) was used to compare estimates between altitude categories.

### Population-level analysis – Mexico

The total cumulative number of COVID-19 cases was significantly higher in Mexican municipalities with a mean elevation ≥2,000-m than in those below 1,500 m (26.0 vs. 19.4 per 100,000; P=0.007) (Figure 1C). The mortality rates were also higher in municipalities located at ≥2,000 m than in those below 1,500 m (5.3 vs. 3.9 per 100,000; P<0.001) (Figure 1D).

### Individual-level analysis – Mexico

Overall, COVID-19 patients living at ≥2,000 m were only marginally older than those living below 1,500 m. However, endotracheal intubation was considerably more common in those living at ≥2,000 m. Likewise, pneumonia and COPD were more common above 2,000 m, whereas hypertension and diabetes were less common (Table 3). Among fatal cases, endotracheal intubation was considerably more common in those living at ≥2,000 m. Likewise, male sex, pneumonia, and COPD were more common above 2,000 m. In contrast, hypertension, cardiovascular disease, and diabetes were less common above 2,000 m (Supplementary Appendix Table S1). Characteristics of COVID-19 cases by survival status are shown in Supplementary Appendix Table S2.

**Table 3.** Characteristics of Mexican subjects with COVID-19.

|  |  | All | Group 1<br>(<1,500 m) | Group 2<br>(1,500-1,999 m) | Group 3<br>(≥2,000 m) | P value*<br>(Group 2 vs.<br>Group 1) | P value*<br>(Group 3 vs.<br>Group 1) |
| --- | --- | --- | --- | --- | --- | --- | --- |
| n |  | 40,168 | 16,858 | 3,209 | 20,101 |  |  |
| Age, yr |  |  |  |  |  | 0.21 | 0.01 |
|  | Mean (SD) | 46.9 (15.7) | 46.8 (16.0) | 46.2 (17.1) | 47.0 (15.3) |  |  |
|  | Median | 46 | 46 | 45 | 46 |  |  |
|  | IQR | 35–57 | 35–57 | 33–58 | 36–57 |  |  |
| Male sex, % |  | 58.2 | 58.4 | 56.9 | 58.2 | -- | -- |
| Intubated, % |  | 4.0 | 3.3 | 4.2 | 4.6 | 0.05 | <0.001 |
| Admitted to ICU, % |  | 3.9 | 4.6 | 3.9 | 3.4 | 0.34 | <0.001 |
| Pneumonia, % |  | 30.2 | 28.1 | 25.7 | 32.6 | 0.018 | <0.001 |
| **Comorbidities** |  |  |  |  |  |  |  |
|  | Hypertension | 21.9 | 24.7 | 19.5 | 19.9 | <0.001 | <0.001 |
|  | Cardiovascular disease | 2.8 | 2.9 | 2.3 | 2.7 | -- | -- |
|  | Obesity | 21.1 | 21.0 | 18.8 | 21.6 | 0.018 | 0.50 |
|  | Diabetes | 18.8 | 19.6 | 16.5 | 18.4 | <0.001 | 0.009 |
|  | Chronic kidney disease | 2.5 | 2.3 | 2.6 | 2.6 | -- | -- |
|  | COPD | 2.3 | 2.0 | 3.1 | 2.5 | <0.001 | 0.004 |
| Time from symptoms onset to<br>hospital admission, days |  |  |  |  |  |  |  |
|  | Among survivors |  |  |  |  | <0.001 | 0.006 |
|  | Mean (SD) | 4.2 (3.3) | 4.2 (3.3) | 3.6 (2.8) | 4.2 (3.4) |  |  |
|  | Median | 4 | 4 | 3 | 4 |  |  |
|  | IQR | 2–6 | 2–6 | 2–5 | 2–6 |  |  |
|  | Among deceased |  |  |  |  | <0.001 | 0.032 |
|  | Mean (SD) | 4.2 (3.3) | 4.3 (3.2) | 3.4 (2.7) | 4.2 (3.4) |  |  |
|  | Median | 4 | 4 | 3 | 4 |  |  |
|  | IQR | 2–6 | 2–6 | 1–5 | 2–6 |  |  |

COPD, chronic obstructive pulmonary disease; ICU, intensive care unit; IQR, interquartile range.
\* P values obtained with the use of the Dunn's (rank-sum) test with Bonferroni adjustment.

We found a significant interaction between age and altitude and between sex and altitude on the association of altitude with pneumonia, requirement for endotracheal intubation, and mortality. Thus, we performed separate regression models for patients younger than 65 years and for those 65 years of age and older. Likewise, we performed separate regression models for women and men.

Among patients younger than 65 years, those who resided at ≥2,000 m had 24% higher risk of pneumonia compared with those who resided below 1,500 m (adjusted IRR: 1.24; 95% confidence interval (CI), 1.00-1.53; P=0.044) adjusting for age, sex, pre-existing comorbidities, and population density of residence location (Figure 2A). The requirement for endotracheal intubation was 78% higher at ≥2,000 m compared with <1,500 m (adjusted IRR: 1.78; 95% CI, 1.15-2.76; P=0.010) (Figure 2B). Likewise, the risk of death attributed to COVID-19 was 36% higher at ≥2,000 m compared with <1,500 m (adjusted IRR: 1.36; 95% CI, 1.05-1.78; P=0.022) (Figure 2C). Among patients 65 years of age and older, we found no differences in the risk of pneumonia (P=0.79), the requirement for endotracheal intubation (P=0.12), or the risk of death (P=0.11) between those who resided at ≥2,000 m and those who resided below 1,500 m (Figure 2).

**Figure 2.**
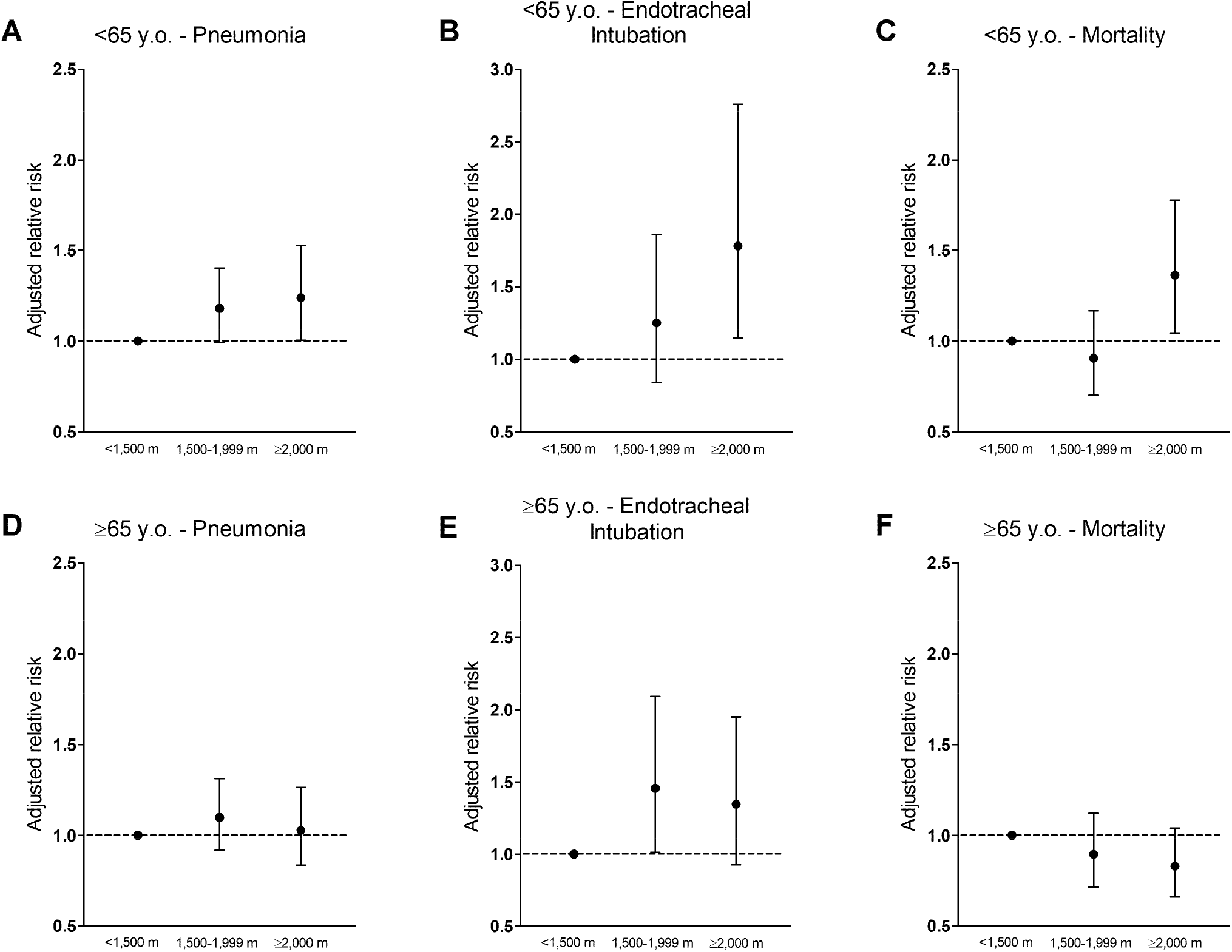
Association between COVID-19 outcomes and altitude categories in Mexican subjects with COVID-19 (n=40,168). Upper panels show the adjusted relative risk (incidence rate ratio, IRR) of pneumonia (A), requirement for endotracheal intubation (B), and death (C) in patients under 65 years of age. Lower panels show the adjusted relative risk of pneumonia (D), requirement for endotracheal intubation (E), and death (F) in patients 65 years of age and older. Estimates were adjusted for age, sex, medical history of diabetes, chronic obstructive pulmonary disease, cardiovascular disease, hypertension, obesity, and chronic kidney disease, and population density of residence location. Vertical lines represent 95% confidence intervals. Altitude <1,500 m represents the reference category (IRR=1.00).

Among women, we found no differences in the risk of pneumonia (P=0.56), the requirement for endotracheal intubation (P=0.10), or the risk of death (P=0.20) between low and high altitude (Supplementary Appendix Figure S1). In contrast, among men, those who resided at ≥2,000 m had 28% higher risk of pneumonia compared with those who resided below 1,500 m (adjusted IRR: 1.28; 95% confidence interval (CI), 1.05-1.56; P=0.016). The requirement for endotracheal intubation was 56% higher at ≥2,000 m compared with <1,500 m (adjusted IRR: 1.56; 95% CI, 1.05-2.31; P=0.028). Likewise, the risk of death attributed to COVID-19 was 31% higher at ≥2,000 m compared with <1,500 m (adjusted IRR: 1.31; 95% CI, 1.03-1.66; P=0.025) (Supplementary Appendix Figure S1).

## DISCUSSION

We found a higher cumulative incidence of COVID-19 cases and higher mortality rates attributed to COVID-19 in populations with a mean altitude ≥2,000-m compared with those with a mean altitude below 1,500 m, both in the U.S. and in Mexico (Figure 1). The differences in the cumulative incidence rates between low- and high-altitude counties may not be seen when the analysis is performed by states as geographical elevation will lose resolution. Because we excluded counties and municipalities with zero deaths in our population-level analyses, our estimates do not represent national estimates of the total number of COVID-19 cases.

Our regression analyses suggest that COVID-19 patients younger than 65 years who live above 2,000 m have a 36% higher adjusted relative risk of death compared with those who live below 1,500 m (Figure 2). Likewise, COVID-19 patients younger than 65 years have a more severe clinical manifestation above 2,000 m, as indicated by a higher requirement for endotracheal intubation and a higher risk of pneumonia. This was not seen in older COVID-19 patients. Men, but not women, also have a 31% higher adjusted relative risk of death and a higher risk of severe clinical manifestation above 2,000 m. It is unclear why the association between altitude and COVID-19 outcomes was significant in the younger population and in men only. This aspect requires further investigation.

The findings of the present study must be interpreted cautiously. Severe hypoxemia and coagulopathy are more common in more severe cases of COVID-19 (Chen et al., 2020; Connors and Levy, 2020). Since chronic environmental hypoxia may aggravate lung disease (Stream et al., 2009) and promote hypercoagulability (Kicken et al., 2018), it is plausible that high altitude hypoxia could contribute to the higher COVID-19 mortality and the severity of COVID-19 in some susceptible individuals, as suggested by our findings. However, our data cannot prove causality. Thus, other possible explanations should also be considered.

Certainly, possible differences in the number of imported cases (e.g. ski tourists, new migrants), population density, and public containment measures across regions, could explain, at least in part, the higher cumulative incidence of COVID-19 cases in high altitude populations. Anecdotal reports of a number of tourists with COVID-19 in Colorado ski resorts suggest that imported cases could represent a confounder in our estimates of COVID-19 cases in the U.S. but would be less relevant in Mexico. However, these factors probably would play a less important role in explaining the higher COVID-19 mortality and the severity of the disease at higher elevations.

Numerous factors are linked to COVID-19 mortality including old age, pre-existing comorbidities, and inadequate healthcare resources (Vincent and Taccone, 2020), all of which could explain the higher mortality above 2,000 m. A strength of our study is that we used a mixed-model regression analysis in a large population of COVID-19 patients to examine the association between altitude and COVID-19 outcomes adjusting for age, sex, and major pre-existing comorbidities, while controlling for nested data. However, we cannot rule out the possible contribution of other unaccounted factors including other comorbidities (e.g. coagulopathies, cancer, immunodeficiency) and ethnic/genetic differences. Additional factors to be considered are altitude-related environmental factors including ambient temperature, air pollution, radiation, and humidity, all of which have been associated with the transmission of SARS-CoV2 (Liu et al., 2020). Our regression model was also adjusted for population density but not for healthcare resources (data unavailable). Information on migration status was also limited in our study population. Among those cases with known migration status (n=139), the proportion of migrants above 2,000 m was less than half of that below 1,500 m.

A major limitation of the present study include possible misreport of COVID-19 cases and deaths. Underreporting of COVID-19 is a global problem (Krantz and Rao, 2020) as the number of cases largely depend on the number of tests performed and the type of test used. This can introduce bias when comparing incidence rates across populations and overestimate or underestimate the total number of deaths attributed to COVID-19. Likewise, it is possible that the number of reported deaths attributed to COVID-19 does not accurately represent the total of fatal cases. Deaths occurring in nursing homes or private residences could be underreported.

## CONCLUSION

In the U.S. and Mexico, populations residing above 2,000 m have a higher total cumulative number of COVID-19 cases and a higher mortality rate attributed to COVID-19 than those residing below 1500 m. Among Mexican subjects with COVID-19, altitude is associated with COVID-19 mortality in men younger than 65 years. Our findings provide new information calling for careful re-examination of public health policies on COVID-19 prevention and deployment of healthcare resources for COVID-19 treatment to high-altitude populations.

## Data Availability

All data are publicly available online.

https://www.newsbreak.com/

https://www.gob.mx/salud

## ACKNOWLEDGMENTS

We thank Newsbreak.com and the Secretary of Health of the Government of Mexico for providing free access to data on COVID-19 cases and deaths.

